# Receipt of anti-SARS-CoV-2 pharmacotherapies among U.S. Veterans with mild to moderate COVID-19, January-February 2022

**DOI:** 10.1101/2022.06.22.22276782

**Authors:** Kristina L. Bajema, Xiao Qing Wang, Denise M. Hynes, Mazhgan Rowneki, Alex Hickok, Francesca Cunningham, Amy Bohnert, Edward J. Boyko, Theodore J. Iwashyna, Matthew L. Maciejewski, Elizabeth M. Viglianti, Elani Streja, Lei Yan, Mihaela Aslan, Grant D. Huang, George N. Ioannou

## Abstract

**Background:** Older adults and persons with medical co-morbidities are at increased risk for severe COVID-19. Several pharmacotherapies demonstrated to reduce the risk of COVID-19-related hospitalization and death have been authorized for use. We describe factors associated with receipt of outpatient COVID-19 pharmacotherapies in the Veterans Health Administration.

**Methods:** We conducted a retrospective cohort study among Veterans with risk factors for severe COVID-19 who tested positive for SARS-CoV-2 during January and February 2022. We compared receipt of any COVID-19 pharmacotherapy, including sotrovimab, nirmatrelvir plus ritonavir, molnupiravir, or remdesivir versus no antiviral or monoclonal antibody treatment according to demographic characteristics, place of residence, underlying medical conditions, and COVID-19 vaccination using multivariable logistic regression.

**Results:** During January and February 2022, 16,546 courses of sotrovimab, nirmatrelvir, and molnupiravir were allocated across the Veterans Health Administration. Among 111,717 Veterans testing positive for SARS-CoV-2, 4,233 (3.8%) received any COVID-19 pharmacotherapy, including 2,870 of 92,396 (3.1%) in January and 1,363 of 19,321 (7.1%) in February. Among a subset of 56,206 Veterans with documented COVID-19-related symptoms in the 30 days preceding positive SARS-CoV-2 test, 3,079 of 53,206 (5.5%) received any COVID-19 pharmacotherapy. Untreated Veterans had a median age of 60 years (interquartile range [IQR] 46-71 years) and median 3 underlying medical conditions (IQR 2-5). Veterans receiving any treatment were more likely to be older (adjusted odds ratio [aOR] 1.66, 95% confidence interval [CI] 1.52-1.80, 65-74 versus 50-64 years; aOR 1.67, 95% CI 1.53-1.84 ≥75 versus 50-64 years) and have a higher number of underlying conditions (aOR 1.63, 95% CI 1.48-1.79, 3-4 versus 1-2 conditions; aOR 2.17, 95% CI 1.98-2.39, ≥5 versus 1-2 conditions). Persons of Black versus White race (aOR 0.65, 95% CI 0.60-0.72) and well as persons of Hispanic ethnicity (aOR 0.88, 95% CI 0.77-0.99) were less likely to receive treatment.

**Conclusions and Relevance:** Although supply of outpatient COVID-19 pharmacotherapies during January and February 2022 was limited, prescription of these pharmacotherapies was underutilized, consistent with early national patterns in dispensing. Racial and ethnic minorities were less likely to receive any pharmacotherapy.

## Introduction

During January 2022 when the incidence of coronavirus disease 2019 (COVID-19) in the United States was at its highest, 82% of intensive care unit hospital beds were occupied, and nearly 900,000 COVID-19-related deaths had occurred since the pandemic began.^1,2^ Older adults and persons with underlying medical conditions such as chronic kidney disease, diabetes, and obesity are at increased risk for severe outcomes including hospitalization or death.^3^ Several neutralizing monoclonal antibodies and antivirals directed at severe acute respiratory syndrome coronavirus 2 (SARS-CoV-2) have received U.S. Food and Drug Administration Emergency Use Authorization (FDA EUA) for treatment of persons with mild to moderate COVID-19 who are at high risk for progression to severe disease.^4-8^ Most recently, these include EUA for nirmatrelvir boosted with ritonavir (nirmatrelvir) and molnupiravir in late December 2021 and remdesivir for outpatient use in January 2022. Although these therapies have been demonstrated in clinical trials to be effective in reducing the short-term risk of hospitalization or death,^9-12^ early limited drug supply, the requirement for prompt recognition of symptomatic disease and linkage to treatment, logistical barriers to administration, and the need for provider and public awareness of therapeutic options have hampered widespread use.^13,14^ To date, utilization of outpatient SARS-CoV-2 pharmacotherapies in the U.S. has not been well-described.

The Veterans Health Administration (VHA) is the largest integrated health care system in the U.S., providing care to over 9 million Veterans at 171 VA Medical Centers (VAMCs) and 1,112 outpatient sites of care.^15^ Within the VHA, COVID-19 pharmacotherapies allocated by the federal government are distributed across 156 VA pharmacies by the VA Pharmacy Benefits Management Services (PBM). This national distribution system serving a population with a majority of older adults with a high burden of underlying conditions who are frequently at increased risk for severe COVID-19^16^ provides a unique opportunity to evaluate how these therapies have been allocated to at-risk patients infected with SARS-CoV-2, including among minority groups for whom reach of novel pharmacotherapies in the general U.S. population is often unequal.^17^ Thus, we sought to describe rates and factors associated with prescription of outpatient COVID-19 pharmacotherapies during January and February 2022 when sotrovimab, a monoclonal antibody active against circulating Omicron SARS-CoV-2 variants at the time and antivirals nirmatrelvir, molnupiravir, and remdesivir were authorized for use.^8^

## Methods

### Study setting and data sources

We used VA’s COVID-19 Shared Data Resource (CSDR),^18^ supported by the VA Informatics and Computing Infrastructure (VINCI) which integrates multiple data sources to provide patient-level COVID-19-related information on VA enrollees. CSDR includes information on first laboratory-confirmed SARS-CoV-2 tests (either by nucleic acid amplification or antigen testing) within the VA system as well as tests performed outside the VA but documented in VA clinical records. Positive tests are identified by the VA National Surveillance Tool^19^ and provisioned to the CSDR to support national VA research and operational needs. We also used the VA Corporate Data Warehouse (CDW), a nationally linked database of the VA electronic health records system (EHR), which contains medical and administrative data as well as pharmacy records. These data were supplemented with detailed claims data (primarily receipt of COVID-19 monoclonal antibodies) from the VA Community Care program, which coordinates and reimburses local care provided outside the VA. We included linked data from the Centers for Medicare and Medicaid Services (CMS), provisioned by the VA Information Resource Center (VIReC), to enhance capture of information on COVID-19 vaccination occurring in non-VA settings. We also integrated data from PBM surveillance on the allocation of sotrovimab, nirmatrelvir, and molnupiravir across VA pharmacies. Remdesivir was approved by FDA in October 2020 for hospitalized patients with COVID-19 and therefore not part of the national PBM distribution of EUA pharmacotherapies.^20^

### Study population and baseline characteristics

We identified Veterans aged 18 years or older with a first positive laboratory-confirmed SARS-CoV-2 test in CSDR between January 1-February 28, 2022. We limited the study population to VA enrollees with an inpatient or outpatient encounter in the VA health care system in the 18 months preceding January 1, 2022, who were not hospitalized on or before the date of positive SARS-CoV-2 test (and hence considered to have mild to moderate COVID-19). We further limited test-positive patients to Veterans with at least one risk factor for progression to severe COVID-19, including hospitalization or death, as defined by the U.S. Food and Drug Administration (FDA) and Centers for Disease Control and Prevention (CDC, Supplementary Table 1).^3-5,21^

Using the date of the positive SARS-CoV-2 test as the index date, we ascertained baseline demographic characteristics and most recent ZIP code of residence documented in the year prior to the index date. The ZIP code was used to determine Veterans Integrated Services Network [VISN], rurality of residence based on the Rural-Urban Commuting Areas [RUCA] system, and distance to the nearest VAMC allocated COVID-19 pharmacotherapies).^22,23^ We also determined COVID-19 vaccination status (Supplemental Methods), current or former smoking, any alcohol or other substance dependence in the two years prior, underlying medical conditions documented within the last two years (Supplementary Table 1), receipt of immunosuppressive medications or cancer therapies filled within the last 90 days for most medications or one year for B-cell depleting therapies (Supplementary Table 2), and National Institutes of Health (NIH) tier of prioritization for anti-SARS-CoV-2 therapies.^24^ Finally, we ascertained 15 pre-specified COVID-19-related symptoms present on the index date or within the preceding 30 days. These symptoms were extracted from EHR ICD-10 codes, vital signs (temperature), and VINCI-generated natural language processing of admission diagnoses, COVID-19 symptom screening questionnaires, and relevant clinical notes.

### COVID-19 Pharmacotherapies

We determined receipt of four FDA authorized therapies in use between January 1-February 28, 2022, which included sotrovimab, nirmatrelvir, molnupiravir, and remdesivir as captured by prescriptions within the VA as well as VA Community Care claims for reimbursed care provided outside of the VA. To distinguish outpatient from inpatient remdesivir use, we reviewed the date of first remdesivir dose relative to the date of hospital admission. Veterans who were not hospitalized after the index date, who received their first dose of remdesivir prior to any hospitalization, or who received their first dose while in the hospital where the length of stay lasted less than one day were classified as having received outpatient remdesivir.

### Statistical analysis

Among Veterans included in our study, we described sociodemographic and clinical characteristics, stratified by receipt of sotrovimab, nirmatrelvir, molnupiravir, remdesivir, or no antiviral or monoclonal antibody (‘no treatment’). Among treated Veterans, we also describe the relative proportion of each of the four pharmacotherapies prescribed by VISN. We conducted binomial logistic regression to estimate odds ratios for receipt of any pharmacotherapy versus no treatment as well as multinomial and binomial logistic regression to estimate odds ratios for receipt of sotrovimab, nirmatrelvir, molnupiravir, or remdesivir versus no treatment. Base models for each factor of interest were adjusted for age, sex, and race/ethnicity. Additional covariates identified *a priori* as potential confounders based on clinical and organizational knowledge (VISN, rural residence, underlying conditions, tobacco, alcohol, substance use) were included for each comparison if they changed the adjusted odds ratio (aOR) by ≥5% when added individually to each base model. Final models were limited to Veterans with complete data for all included covariates. Odds ratios were compared using 95% confidence intervals (CIs). Analyses were conducted using SAS Enterprise Guide 8.2 (SAS Institute Inc). The study was approved by the VA Puget Sound and VA Portland Institutional Review Boards.

## Results

### Descriptive Results

During January and February 2022, there were an estimated 16,546 total courses of sotrovimab (n=3,770), nirmatrelvir (n=5,220), and molnupiravir (n=7,556) distributed across the VHA and 111,717 VA enrollees with a first positive SARS-CoV-2 test who were potentially eligible to receive therapy for mild to moderate COVID-19 (Figure 1), of whom 4,233 (3.8%) received any pharmacotherapy within the VA or through VA Community Care. This included 994 (0.9%) persons who received sotrovimab, 1,710 (1.5%) who received nirmatrelvir, 921 (0.8%) who received molnupiravir, 608 (0.5%) who received remdesivir and 107,484 (96.2%) who were not treated with anti-SARS-CoV-2 pharmacotherapies within the VA or through VA Community Care (Table 1).

**Figure 1.**
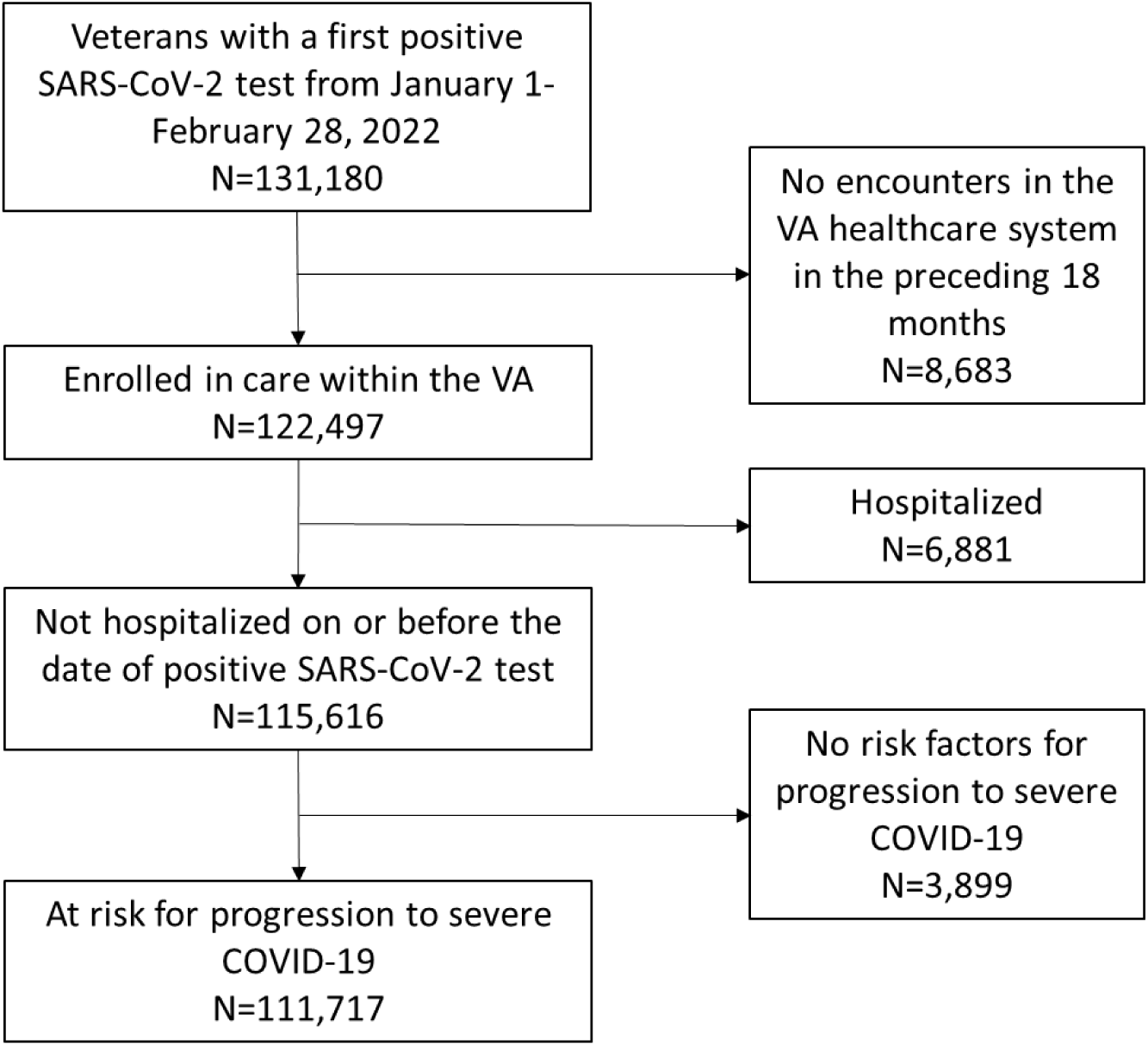
Veterans with a first positive SARS-CoV-2 test from January 1-February 28, 2022 included in the study. Potentially eligible Veterans had an inpatient or outpatient encounter in the VA health care system in the 18 months preceding January 1, 2022, were not hospitalized on or before the date of positive SARS-CoV-2 test, and had at least one risk factor for progression to severe COVID-19.^3-5,21^

**Table 1.** Characteristics of Veterans testing positive for SARS-CoV-2 by receipt of outpatient COVID-19 pharmacotherapy, January-February 2022^a^

| Characteristic | All Veterans | No Treatment | Any Treatment* | Sotrovimab | Nirmatrelvir and ritonavir | Molnupiravir | Remdesivir |
| --- | --- | --- | --- | --- | --- | --- | --- |
| Total<sup>a</sup> | 111,717 | 107,484 | 4,233 | 994 | 1,710 | 921 | 608 |
| Age, years, median [IQR] | 60 [46,72] | 60 [46,71] | 68 [58,74] | 71 [63,75] | 66 [54,74] | 68 [57,75] | 68 [59, 75] |
| Age group, N (%) |  |  |  |  |  |  |  |
| 18-49 | 33,206 (30) | 32,646 (30) | 560 (13) | 86 (9) | 293 (17) | 113 (12) | 68 (11) |
| 50-64 | 33,976 (30) | 32,849 (31) | 1,127 (27) | 193 (19) | 501 (29) | 254 (28) | 179 (29) |
| 65-74 | 26,971 (24) | 25,464 (24) | 1,507 (36) | 416 (42) | 566 (33) | 323 (35) | 202 (33) |
| ≥75 | 17,564 (16) | 16,525 (15) | 1,039 (24) | 299 (30) | 350 (20) | 231 (25) | 159 (26) |
| Male sex, N (%) | 96,482 (86) | 92,648 (86) | 3,834 (91) | 911 (92) | 1,527 (89) | 836 (91) | 560 (92) |
| Race, N (%)<sup>b</sup> |  |  |  |  |  |  |  |
| White | 75,973 (68) | 72,726 (68) | 3,247 (77) | 769 (77) | 1,318 (77) | 721 (78) | 439 (72) |
| Black | 23,362 (21) | 22,740 (21) | 622 (15) | 127 (13) | 252 (15) | 137 (15) | 106 (17) |
| Other | 3,896 (3) | 3,783 (4) | 113 (3) | 34 (3) | 43 (2) | 20 (2) | 16 (3) |
| Ethnicity, N (%) |  |  |  |  |  |  |  |
| Not Hispanic | 95,607 (86) | 91,870 (85) | 3,737 (88) | 888 (89) | 1,492 (87) | 828 (90) | 529 (87) |
| Hispanic | 10,740 (10) | 10,414 (10) | 326 (8) | 70 (7) | 156 (9) | 54 (6) | 46 (8) |
| Rurality, N (%)<sup>d</sup> |  |  |  |  |  |  |  |
| Urban | 94,217 (84) | 90,832 (85) | 3,385 (80) | 775 (78) | 1,396 (82) | 717 (78) | 497 (82) |
| Rural | 17,500 (16) | 16,652 (15) | 848 (20) | 219 (22) | 314 (18) | 204 (22) | 111 (18) |
| Region, N (%)<sup>e</sup> |  |  |  |  |  |  |  |
| West | 27,931 (25) | 27,012 (25) | 919 (22) | 209 (21) | 366 (21) | 224 (24) | 120 (20) |
| Midwest | 20,451 (18) | 19,522 (18) | 929 (22) | 256 (26) | 422 (25) | 208 (23) | 43 (7) |
| Northeast | 15,432 (14) | 14,642 (14) | 790 (19) | 208 (21) | 339 (20) | 97 (11) | 146 (24) |
| South | 47,903 (43) | 46,308 (43) | 1,595 (37) | 321 (32) | 583 (34) | 392 (43) | 299 (49) |
| Distance from place of residence to nearest VAMC allocated the respective product, miles, median [IQR]<sup>f</sup> | 18 [8,39] | 18 [8, 39] | 17 [8,34] | 20 [8,41] | 16 [7,31] | 16 [7,32] | N/A |
| Month of positive test, N (%) |  |  |  |  |  |  |  |
| January | 92,396 (83) | 89,526 (83) | 2,870 (68) | 651 (65) | 1,064 (62) | 647 (70) | 508 (84) |
| February | 19,321 (17) | 17,958 (17) | 1,363 (32) | 343 (35) | 646 (38) | 274 (30) | 100 (16) |
| ≥1 symptom in the 30 days prior to positive SARS-CoV-2 test, N (%) | 56,285 (50) | 53,206 (50) | 3,079 (73) | 707 (71) | 1,228 (72) | 696 (76) | 448 (74) |
| Vaccination status, N (%)<sup>g</sup> |  |  |  |  |  |  |  |
| No vaccination | 34,258 (31) | 33,108 (31) | 1,150 (27) | 276 (28) | 500 (29) | 212 (23) | 162 (27) |
| Partial vaccination | 4,369 (4) | 4,135 (4) | 234 (6) | 69 (7) | 84 (5) | 37 (4) | 44 (7) |
| Primary vaccination | 46,079 (41) | 44,482 (41) | 1,597 (38) | 402 (40) | 616 (36) | 361 (39) | 218 (36) |
| Booster vaccination | 26,980 (24) | 25,728 (24) | 1,252 (29) | 247 (25) | 510 (30) | 311 (34) | 184 (30) |
| Other | 31 (0) | 31 (0) | 0 (0) | 0 (0) | 0 (0) | 0 (0) | 0 (0) |
| NIH Tier, N (%)<sup>h</sup> |  |  |  |  |  |  |  |
| 1 | 14,987 (13) | 13,913 (13) | 1,074 (25) | 385 (39) | 360 (21) | 161 (17) | 168 (28) |
| 2 | 25,559 (23) | 24,985 (23) | 574 (14) | 83 (8) | 294 (17) | 118 (13) | 79 (13) |
| 3 | 31,995 (29) | 30,313 (28) | 1,682 (40) | 398 (40) | 640 (37) | 411 (45) | 233 (38) |
| 4 | 39,148 (35) | 38,245 (36) | 903 (21) | 128 (13) | 416 (24) | 231 (25) | 128 (21) |
| Smoking, N (%)<sup>i</sup> |  |  |  |  |  |  |  |
| Never | 43,676 (39) | 42,085 (39) | 1,591 (38) | 355 (36) | 673 (39) | 329 (35) | 234 (38) |
| Former | 44,506 (40) | 42,626 (40) | 1,880 (44) | 485 (49) | 720 (42) | 411 (45) | 264 (43) |
| Current | 19,057 (17) | 18,454 (17) | 603 (14) | 112 (11) | 255 (15) | 156 (17) | 80 (13) |
| Alcohol dependence, N (%) |  |  |  |  |  |  |  |
| Never | 86,379 (77) | 82,856 (77) | 3,523 (83) | 845 (85) | 1,414 (83) | 762 (83) | 502 (83) |
| Ever | 25,338 (23) | 24,628 (23) | 710 (17) | 149 (15) | 296 (17) | 159 (17) | 106 (17) |
| Substance dependence, N (%) |  |  |  |  |  |  |  |
| Never | 105,845 (95) | 101,778 (95) | 4,067 (96) | 965 (97) | 1,651 (97) | 881 (96) | 570 (94) |
| Ever | 5,872 (5) | 5,706 (5) | 166 (4) | 29 (3) | 59 (3) | 40 (4) | 38 (6) |
| Number of underlying conditions, median [IQR] | 3 [2,5] | 3 [2,5] | 4 [3,14] | 5 [3,15] | 4 [2,7] | 4 [3,14] | 5 [3,15] |
| Charlson Comorbidity Index, mean | 1.5 | 1.5 | 2.4 | 3.2 | 1.9 | 2.4 | 2.7 |
| Underlying condition, N (%) |  |  |  |  |  |  |  |
| Obese (body mass index $\geq 30$ kg/m<sup>2</sup>) | 56,708 (51) | 54,535 (51) | 2,173 (51) | 505 (51) | 886 (52) | 497 (54) | 285 (47) |
| Chronic kidney disease | 13,594 (12) | 12,749 (12) | 845 (20) | 319 (32) | 216 (13) | 192 (21) | 118 (19) |
| Diabetes | 33,543 (30) | 31,718 (30) | 1,825 (43) | 508 (51) | 664 (39) | 386 (42) | 267 (44) |
| Immuno-suppressive medications or cancer therapies<sup>j</sup> | 4,760 (4) | 4,242 (4) | 518 (12) | 211 (21) | 150 (9) | 63 (7) | 94 (15) |
| Cancer | 20,150 (18) | 18,967 (18) | 1,183 (28) | 350 (35) | 448 (26) | 229 (25) | 156 (26) |
| Cardiovascular disease | 67,149 (60) | 63,949 (60) | 3,200 (76) | 829 (83) | 1,193 (70) | 712 (77) | 466 (77) |
| Chronic lung disease | 23,096 (21) | 21,817 (20) | 1,279 (30) | 349 (35) | 447 (26) | 288 (31) | 195 (32) |
| Dementia | 3,889 (3) | 3,656 (3) | 233 (6) | 54 (5) | 69 (4) | 44 (5) | 66 (11) |
| Cerebrovascular disease | 5,579 (5) | 5,236 (5) | 343 (8) | 91 (9) | 93 (5) | 88 (10) | 71 (12) |
| Chronic liver disease | 9,642 (9) | 9,191 (9) | 451 (11) | 125 (13) | 160 (9) | 88 (10) | 78 (13) |
| Mental health conditions<sup>k</sup> | 46,349 (41) | 44,729 (42) | 1,620 (38) | 345 (35) | 622 (36) | 386 (42) | 267 (44) |

<sup>a</sup>Among Veterans enrolled in care, not hospitalized at the time of positive test, and with $\geq 1$ risk factor for severe COVID-19.
<sup>b</sup>Race unknown: 8,486 (8%) in the all Veterans group, 8,235 (8%) in the no treatment group, 251 (6%) in any treatment group, 64 (6%) who received sotrovimab, 97 (6%) who received nirmaltrelvir, 43 (5%) who received molnupiravir, and 47 (8%) who received remdesivir. Other race includes Asian, American Indian or Alaska Native, Native Hawaiian or Pacific Islander.
<sup>c</sup>Ethnicity unknown: 5,370 (5%) in the all patient group, 5,200 (5%) in the no treatment group, 170 (4%) in any treatment group, 36 (4%) who received sotrovimab, 62 (4%) who received nirmaltrelvir, 39 (4%) who received molnupiravir, and 33 (5%) who received remdesivir.
<sup>d</sup>Based on rural-urban commuting area (RUCA) codes.
<sup>e</sup>Regions are based on Veterans Integrated Service Networks (VISNs). West includes VISNs 19-22; Midwest 10,12,15,23; Northeast 1,2,4,5; South 6-9, 16-17.
<sup>f</sup>For the no treatment group, distance is calculated to the nearest facility prescribing any treatment.
<sup>g</sup>See Supplemental Methods.
<sup>h</sup>As defined by NIH: https://www.covid19treatmentguidelines.nih.gov/overview/prioritization-of-therapeutics/?msclkid=9db67596cf2311ec8b6f159cc2c59087. See Supplementary Table 3.
<sup>i</sup>Smoking unknown: 4,478 (4%) in the all patient group, 4,319 (4%) in the no treatment group, 159 (4%) in any treatment group, 42 (4%) who received sotrovimab, 62 (4%) who received nirmaltrelvir, 25 (3%) who received molnupiravir, and 30 (5%) who received remdesivir.
<sup>j</sup>See Supplementary Table 2.
<sup>k</sup>Includes major depressive disorder, bipolar disorder, schizophrenia.
Abbreviations: IQR, interquartile range; NIH, National Institutes of Health

Approximately 3.1% of Veterans diagnosed in January and 7.1% of Veterans diagnosed in February received any of the pharmacotherapies of interest. Among 53,206 Veterans with any COVID-19-related symptoms, 3,079 (5.5%) received any pharmacotherapy, while 1,384 of 37,274 (3.4%) of Veterans who had not completed primary or booster COVID-19 vaccination received any pharmacotherapy. Of the total courses of PBM-distributed EUA pharmacotherapies, a total of 3,625 (21.9%) were prescribed to eligible Veterans, including 26.4% of the available sotrovimab supply, 32.8% of nirmatrelvir, and 12.2% of molnupiravir.

Among untreated Veterans, median age was 60 years (interquartile range [IQR] 46-71 years), 72,726 (68%) were White and 22,740 (21%) Black race, 70,210 (65%) had completed primary or booster COVID-19 vaccination, and median number of underlying conditions was 3 (IQR 2-5, Table 1). Among persons treated with sotrovimab, median age was 71 years (IQR 63-75 years), 767 (77%) were White and 127 (13%) Black race, 649 (65%) had received full or boosted COVID-19 vaccination, and median number of underlying conditions was 5 (IQR 3-15). Among persons treated with nirmatrelvir, median age was 66 years (IQR 54-74 years), 1,318 (77%) were White and 252 (15%) Black race, 1,116 (66%) had received full or boosted COVID-19 vaccination, and median number of underlying conditions was 4 (IQR 2-7). Among persons treated with molnupiravir, median age was 68 years (IQR 57-75 years), 721 (78%) were White and 137 (15%) Black race, 672 (73%) had received full or boosted COVID-19 vaccination, and median number of underlying conditions was 4 (IQR 3-14). Among persons treated with remdesivir, median age was 68 years (IQR 59-75 years), 439 (72%) were White and 106 (17%) Black race, 402 (66%) had received full or boosted COVID-19 vaccination, and median number of underlying conditions was 5 (IQR 3-15).

#### Regression Results

Veterans receiving any of the four treatments were more likely to be older (aOR 1.66, 95% CI 1.52-1.80, 65-74 versus 50-64 years; aOR 1.67, 95% CI 1.53-1.84 ≥75 versus 50-64 years), live in rural areas (aOR 1.18, 95% CI 1.09-1.28), and have a higher number of underlying conditions (aOR 1.63, 95% CI 1.48-1.79, 3-4 versus 1-2 conditions; aOR 2.17, 95% CI 1.98-2.39, ≥5 versus 1-2 conditions, Table 2). Receipt of immunosuppressive medications or cancer treatments was strongly associated with treatment (aOR 3.03, 95% CI 2.74-3.36). Persons of Black versus White race (aOR 0.65, 95% CI 0.60-0.72) and Hispanic ethnicity (aOR 0.88, 95% CI 0.77-0.99) were less likely to receive treatment as were persons with alcohol dependence (aOR 0.79, 95% CI 0.72-0.86). Veterans partially vaccinated against COVID-19 were more likely to receive any treatment compared with unvaccinated Veterans (aOR 1.52, 95% CI 1.30-1.77), whereas there were no significant differences among fully vaccinated (aOR 0.96, 95% CI 0.89-1.04) or boosted persons (aOR 1.06, 95% CI 0.97-1.16). Factors associated with receipt of any treatment were similar when evaluated by individual pharmacotherapies including sotrovimab, nirmatrelvir, molnupiravir, or remdesivir (Supplementary Table 4), or when restricting to symptomatic individuals (Supplementary Table 5).

**Table 2.** Factors associated with receipt of any COVID-19 pharmacotherapy^a^ among Veterans, January-February 2022

| Characteristic | Any pharmacotherapy/total n/N (%)<sup>b</sup> | Any pharmacotherapy Adjusted odds ratio<sup>c</sup> (95% CI) |
| --- | --- | --- |
| Age, years |  |  |
| 18-49 | 507/28,890 (1.8) | 0.51 (0.46-0.56) |
| 50-64 (ref) | 1,030/30,749 (3.4) | - |
| 65-74 | 1,401/24,815 (5.7) | 1.66 (1.52-1.80) |
| ≥75 | 952/16,133 (5.9) | 1.67 (1.53-1.84) |
| Female sex | 359/13,526 (2.7) | 0.97 (0.86-1.09) |
| Race |  |  |
| White (ref) | 3,179/74,093 (4.3) | - |
| Black | 606/22,756 (2.7) | 0.65 (0.6-0.72) |
| Other | 105/3,738 (2.8) | 0.76 (0.62-0.93) |
| Hispanic ethnicity | 272/8,609 (3.2) | 0.88 (0.77-0.99) |
| Rural residence | 791/16,137 (4.9) | 1.18 (1.09-1.28) |
| Region |  |  |
| West (ref) | 822/23,959 (3.4) | - |
| Midwest | 872/18,831 (4.6) | 1.32 (1.20-1.46) |
| Northeast | 732/14,147 (5.2) | 1.51 (1.36-1.67) |
| South | 1,464/43,650 (3.4) | 1.03 (0.94-1.12) |
| COVID-19 vaccination |  |  |
| None (ref) | 1,045/30,671 (3.4) | - |
| Partial | 209/3,917 (5.3) | 1.52 (1.30-1.77) |
| Primary | 1,476/41,418 (3.6) | 0.96 (0.89-1.04) |
| Booster | 1,160/24,557 (4.7) | 1.06 (0.97-1.16) |
| Smoking |  |  |
| Never (ref) | 1,462/39,061 (3.7) | - |
| Former | 1,728/40,274 (4.3) | 0.99 (0.92-1.07) |
| Current | 562/17,286 (3.3) | 0.90 (0.81-0.99) |
| Alcohol dependence | 646/22,658 (2.9) | 0.79 (0.72-0.86) |
| Substance dependence | 154/5,400 (2.9) | 0.90 (0.76-1.06) |
| Number of underlying conditions |  |  |
| 1-2 (ref) | 790/37,502 (2.1) | - |
| 3-4 | 1,222/31,289 (3.9) | 1.63 (1.48-1.79) |
| ≥5 | 1,845/29,852 (6.2) | 2.17 (1.98-2.39) |
| Cancer | 1,102/18,626 (5.9) | 1.38 (1.29-1.49) |
| Cardiovascular disease | 1,790/30,052 (6.0) | 1.52 (1.41-1.63) |
| Chronic kidney disease | 783/12,539 (6.2) | 1.40 (1.29-1.53) |
| Chronic lung disease | 1,558/28,957 (5.4) | 1.39 (1.31-1.49) |
| Diabetes | 1,688/30,708 (5.5) | 1.43 (1.33-1.53) |
| Immunosuppressive or cancer medications | 471/4,361 (10.8) | 3.03 (2.74-3.36) |
| Mental health conditions | 1,497/41,717 (3.6) | 1.13 (1.05-1.21) |
| Obese (body mass index ≥30 kg/m<sup>2</sup>) | 2,002/51,129 (3.9) | 1.19 (1.11-1.27) |

<sup>a</sup>Includes sotrovimab, nirmatrelvir, molnupiravir, and remdesivir.
<sup>b</sup>A total of 4,233 Veterans who received any COVID-19 pharmacotherapy out of 111,717 Veterans testing positive for SARS-CoV-2 were included. Models were limited to Veterans with complete data for all included covariates.
<sup>c</sup>All models adjusted for age, sex, race, and ethnicity.

Among Veterans treated with anti-SARS-CoV-2 pharmacotherapies, the relative proportion of treatments varied across VISNs, from 3-37% for sotrovimab, 30-64% for nirmatrelvir, 6-45% for molnupiravir, and 2-30% for remdesivir (Figure 2).

**Figure 2.**
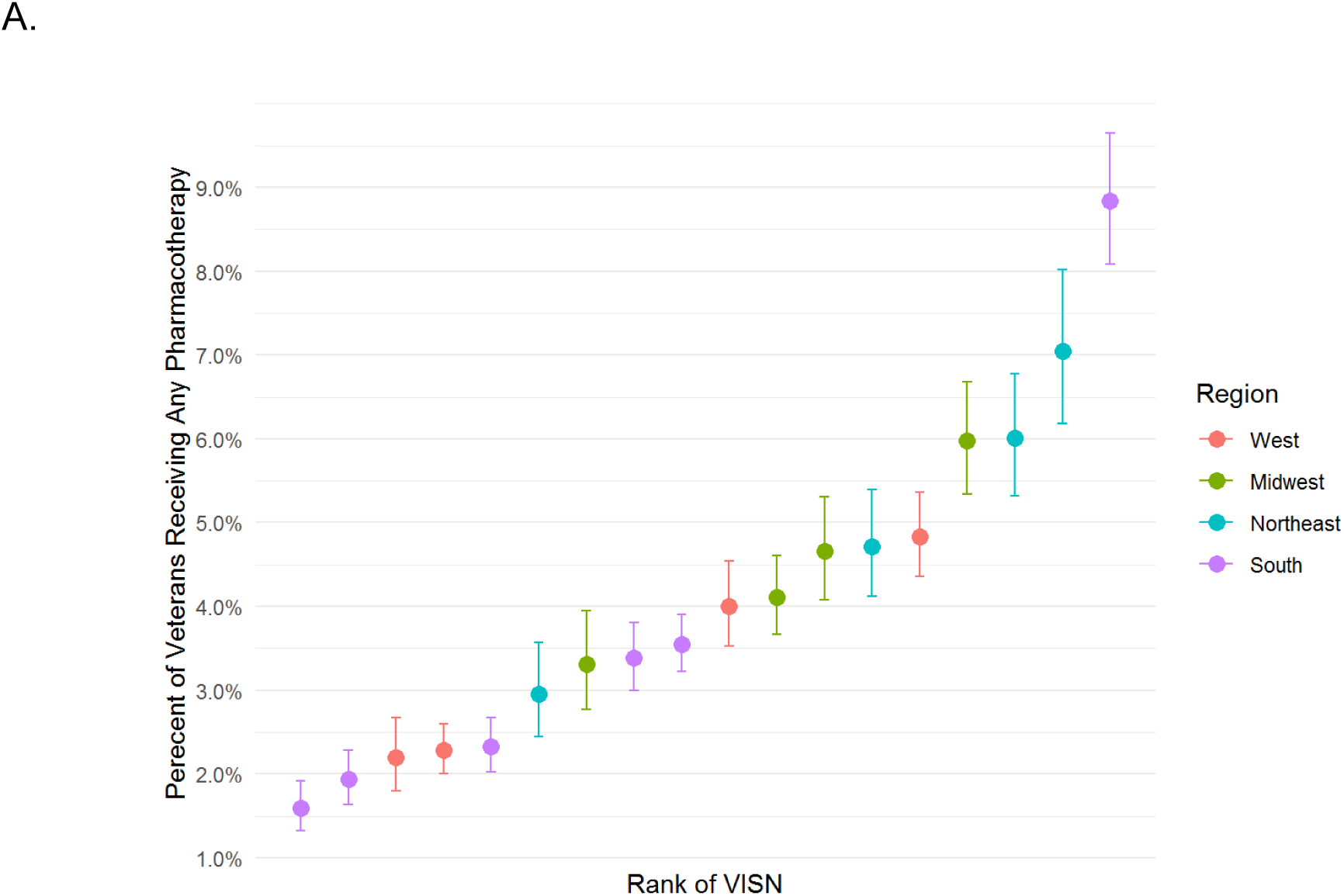
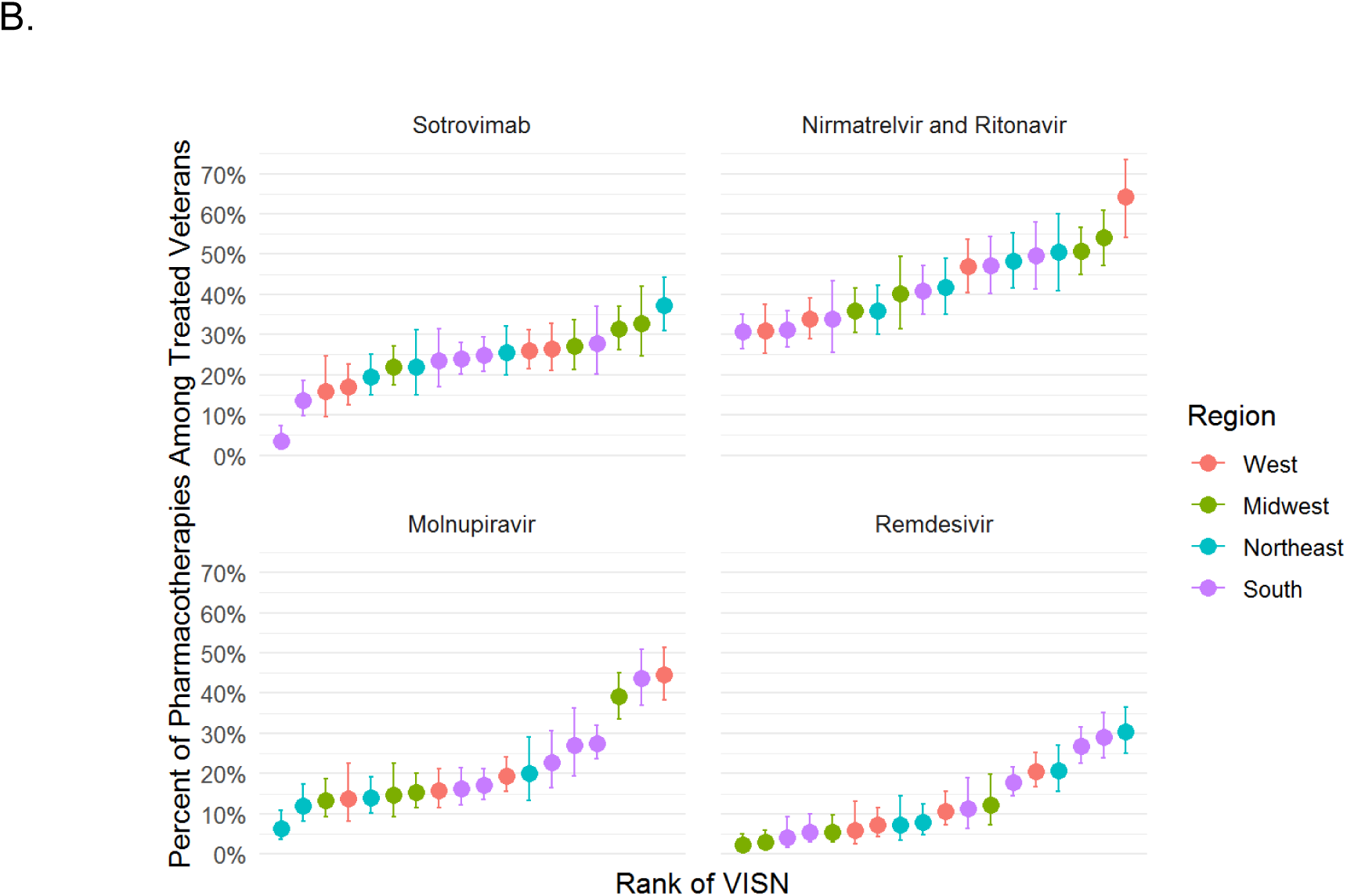
Distribution of COVID-19 pharmacotherapies by Veterans Integrated Services Networks. (A) Percentage of Veterans receiving any COVID-19 pharmacotherapy, (B) Pharmacotherapies prescribed among treated Veterans. Regions are based on VISNs. West includes VISNs 19-22; Midwest 10,12,15,23; Northeast 1,2,4,5; South 6-9, 16-17. Error bars indicate 95% confidence intervals for proportions. Abbreviation: VISN, Veterans Integrated Services Network

## Discussion

In a nationwide study of 111,717 U.S. Veterans seen in the VA healthcare system who tested positive for SARS-CoV-2 in January and February 2022 and were potentially eligible to receive outpatient anti-SARS-CoV-2 pharmacotherapies for mild-to-moderate COVID-19, only 3.8% of Veterans received any outpatient pharmacotherapy, including 5.5% of those with documented COVID-19-related symptoms and 3.4% of those who were unvaccinated or only partially vaccinated. Untreated Veterans included many persons at high risk for progression to severe COVID-19. Persons of Black race and Hispanic ethnicity were less likely to receive treatment while older Veterans with a higher number of underlying conditions were more likely to receive treatment. There were notable geographic differences in the distribution of selected pharmacotherapies.

Prescription of outpatient SARS-CoV-2 pharmacotherapies following FDA EUA of the oral antivirals in December 2021^8^ has not been well-described. As reported here, even among at-risk patients engaged in care in a system with national distribution across 156 sites, utilization remained low. A similarly low uptake of monoclonal antibody treatments during 2020-2021 was described among Medicare beneficiaries as well as patients within a national clinical research network in the U.S.^13,25^ Because the oral antivirals were authorized just before the start of the study period, it is expected for initial uptake to be low and increase with time as clinical familiarity and infrastructures develop; in fact, the proportion of Veterans testing positive for SARS-CoV-2 who were treated more than doubled from 3.1% in January to 7.1% in February. Nonetheless, there was still a relative surplus in drug supply during this period. Low utilization during this early rollout period was not unique to the VA and was also observed in the setting of Department of Health and Human Services-led efforts to widely distribute COVID-19 pharmacotherapies across the U.S,^26,27^ reflecting widespread, complex barriers to optimal use.

Across VISNs, there were notable differences in the percentage of Veterans receiving any COVID-19 pharmacotherapy as well the relative utilization of different pharmacotherapies. Because there was a relative surplus of COVID-19 pharmacotherapies in the VA, circulated on a similar timeline across 156 facilities, this likely reflects differences in local infrastructure, provider education, and perceptions about national treatment availability. Furthermore, in the absence of real-world evidence for the effectiveness of authorized therapies or head-to-head clinical trials, clinical practice has varied. Local and regional differences in patient awareness of therapies, particularly during the period of this study before the launch of the nationwide COVID-19 Test To Treat Initiative,^28^ also likely contributed to variability in prescribing across VISNs.

Differences in treatment by rurality were also observed, with Veterans in rural areas somewhat more likely to receive treatment. Although U.S. Veterans are a highly rural population,^29^ many VA facilities allocated anti-SARS-CoV-2 pharmacotherapies are also located in rural areas. Rural Veterans in our study, in addition to being older, male, and non-Hispanic White, also had more underlying conditions than their urban counterparts (data not shown). Models were adjusted for demographics but not underlying conditions as these are in the causal pathway between rurality and treatment.^30^ Finally, Veterans testing positive for SARS-CoV-2 lived within a relatively close distance of a dispensing facility, and we did not observe a meaningful difference between treated and untreated persons, suggesting that relative to other factors, physical distance may not have been a significant barrier to treatment.

Consistent with findings in other studies, Black Veterans were less likely to receive newly available outpatient treatments for COVID-19; Hispanic Veterans in our study were also slightly less likely to receive treatment, whereas other studies have been mixed.^13,25^ While demographic differences in access to care, including lower use of COVID-19 monoclonal antibody treatments among racial and ethnic minorities have been well-described in non-Veteran populations,^25^ disparities in COVID-19-related care within the VA system have been less pronounced.^16,31^ Possible reasons for observed racial and ethnic differences in treatment of mild to moderate COVID-19 include structural barriers such as limited access to testing, lower awareness of COVID-19 therapies, differences in care-seeking, and lower trust in the healthcare system impacting acceptance of recommended investigational therapies.^14,32,33^

The association between increasing age and higher number of underlying conditions with receipt of pharmacotherapy is consistent with NIH patient prioritization recommendations, which include age, immune status, and clinical risk factors as key determinants for assessing risk for progression to severe COVID-19.^24^ NIH guidance also prioritizes unvaccinated individuals, however we did not observe a consistent pattern between likelihood of treatment and COVID-19 vaccination; despite the early emphasis on treating unvaccinated individuals, we did not observe any differences in the likelihood of treatment in this group compared with fully vaccinated or boosted Veterans. On the other hand, partially vaccinated Veterans were more likely to be treated. These findings may reflect differences in patient behaviors, with Veterans willing to seek vaccination also more likely to accept recommended investigational therapies. Differences in provider practice may also affect observed results; because the demonstrated benefit of COVID-19 pharmacotherapies in clinical trials was among unvaccinated participants, clinicians may have differing perspectives on the potential of these therapies to reduce the risk of severe COVID-19 according to vaccination status. With nearly two-thirds of Veterans in our study fully vaccinated or boosted, real-world evidence for the effectiveness of current used therapies will be essential to inform best practices.

This study has several important limitations. Eligibility for treatment of mild to moderate COVID-19 under FDA EUA requires symptomatic disease, and we were not able to fully ascertain COVID-19-related symptoms. National surveillance is conducted by VA PBM to ensure eligibility among Veterans receiving treatment in the VA; however, determination of symptom eligibility among untreated Veterans in this study is limited to use of natural language processing supplementation of structured data. While symptoms were only captured in 50% of untreated Veterans, this is likely an underascertainment of the true proportion experiencing COVID-19-related symptoms. Nonetheless, even with the study population restricted to documented symptomatic individuals, the percentage of Veterans (5.5%) receiving treatment was still small. While we augmented data from the VA EHR with multiple sources of information, including receipt of COVID-19 monoclonal antibodies through the VA Community Care program, it is still possible that Veterans were treated for COVID-19 outside of the VA. However, the study was conducted during a time when treatments were still very limited. We also restricted the study population to Veterans engaged in VA care to increase the likelihood that treatment would occur within the VA system. While CSDR captures laboratory-based SARS-CoV-2 tests performed in the VA as well as non-VA testing documented in clinical notes, we did not identify all Veterans testing positive for SARS-CoV-2, in particular those who did not report non-VA or home-based testing to their provider. Thus, we may have underestimated the true number of untreated Veterans.

Furthermore, our estimates of prescribed pharmacotherapies do not account for Veterans who were offered but declined treatment. Finally, Veterans are older and have more underlying conditions than persons in the general U.S. population, and care delivery in the VA is very different from non-VA systems, therefore, utilization findings may not be generalizable to other groups.

## Conclusions

In this nationwide study of U.S. Veterans using the VA healthcare system and testing positive for SARS-CoV-2 during January and February 2022, most eligible persons were not prescribed newly available treatments for mild to moderate COVID-19. Demographic, clinical, and facility-level factors were associated with the likelihood of being prescribed treatment. These findings reinforce the need for improved infrastructure and education to support treatment of persons at risk for progression to severe COVID-19. Real world studies of the effectiveness and comparative effectiveness of outpatient COVID-19 pharmacotherapies will also be critical to inform clinical care.

## Data Availability

All data produced in the present work are contained in the manuscript

## Disclaimer

The contents do not represent the views of the U.S. Department of Veterans Affairs or the US Government.

## Additional Contributions

We thank the Biomedical Advanced Research and Development Authority (BARDA) for their support.

## Supplemental Material

### Supplemental Methods

#### COVID-19 Vaccination Status

Veterans were considered unvaccinated if they did not receive any COVID-19 vaccine or received a vaccine dose other than Janssen less than 14 days prior to the first positive SARS-CoV-2 test (index date). Partial vaccination was defined as having received a single mRNA dose (Pfizer-BioNTech or Moderna) alone or in combination with another vaccine other than Janssen ≤14 days prior to the index date or a Janssen (Johnson & Johnson) dose <14 days before the index date. For Veterans not considered to be moderately or severely immunocompromised (i.e., had not recently received immunosuppressive or cancer medications as described in Supplementary Table 2), full vaccination was defined as receipt of both doses of an mRNA vaccine (Pfizer-BioNTech or Moderna) or a single Janssen dose ≥14 days before the index date. Non-immunocompromised Veterans were considered boosted if they either received 1 dose of Janssen followed by another dose of Janssen or mRNA vaccine ≥14 days before the index date or if they received ≥3 vaccine doses (which included at least 1 mRNA dose) ≥7 days before the index date. Moderately or severely immunocompromised Veterans who either received 3 mRNA vaccine doses ≥7 days before the index date or 1 dose of Janssen followed by another dose of Janssen or mRNA vaccine ≥14 days before the index date were considered fully vaccinated.^34^ Immunocompromised Veterans who received ≥4 mRNA doses ≥7 days before the index date were considered boosted.

#### NIH Tiers

In order to categorize vaccination as a binary variable for the National Institutes of Health (NIH) tiers of prioritization for anti-SARS-CoV-2 therapies, we considered unvaccinated and partially vaccinated Veterans as ‘unvaccinated’ for tiers 1 and 2 and fully vaccinated and boosted as ‘vaccinated.’

**Supplementary Table 1.** Risk factors for severe COVID-19^a^

|  |
| --- |
| **Age $\geq 65$ years** |
| **Underlying Medical Conditions** |
| Cancer |
| Cardiovascular disease including cardiomyopathy, chronic rheumatic heart disease, congestive heart failure, coronary artery disease, hypertension, myocardial infarction, peripheral artery disease, pulmonary heart disease |
| Chronic kidney disease including dialysis |
| Chronic liver disease including chronic hepatitis and cirrhosis |
| Chronic lung disease including asthma, chronic obstructive pulmonary disease, emphysema, pulmonary fibrosis |
| Chronic neurologic conditions including epilepsy, Multiple Sclerosis, and Parkinson's Disease |
| Dementia |
| Diabetes |
| HIV |
| Immunosuppressive medications or cancer therapies (see Supplementary Table 2) |
| Mental health conditions including bipolar disorder, major depressive disorder, and schizophrenia |
| Overweight (body mass index 25 to <30 kg/m<sup>2</sup>) or obese (body mass index ≥30 kg/m<sup>2</sup>) or |
| Pregnancy |
| Sickle cell disease |
| Stroke or cerebrovascular disease |
| Thalassemia |
| **Current or former tobacco use** |
| **Substance use** |
| Alcohol dependence |
| Non-alcohol substance dependence |

<sup>a</sup>Age at the time of positive SARS-CoV-2 test or underlying conditions, tobacco, or substance use documented in the two years prior to positive SARS-CoV-2 test.

**Supplementary Table 2.**
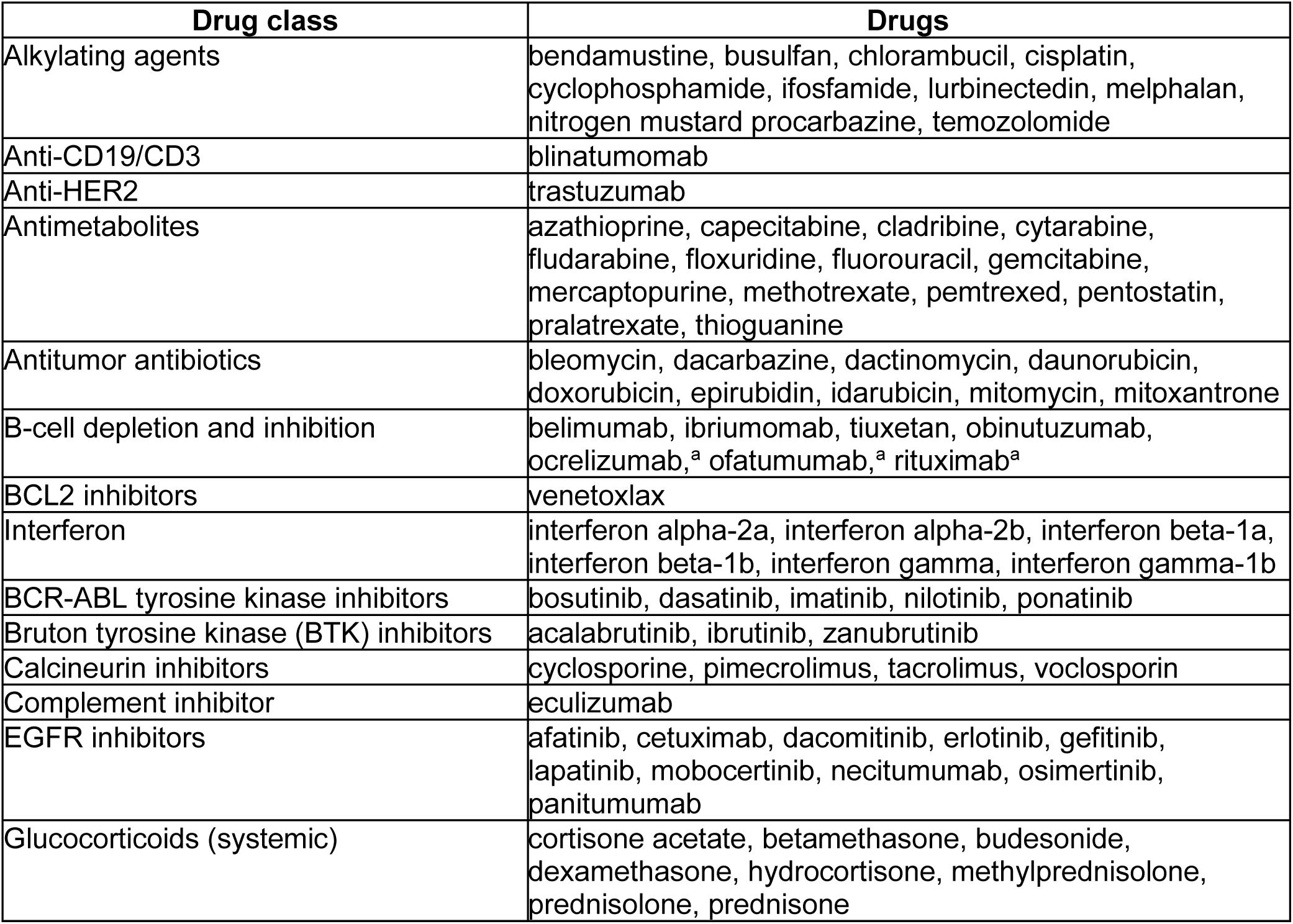
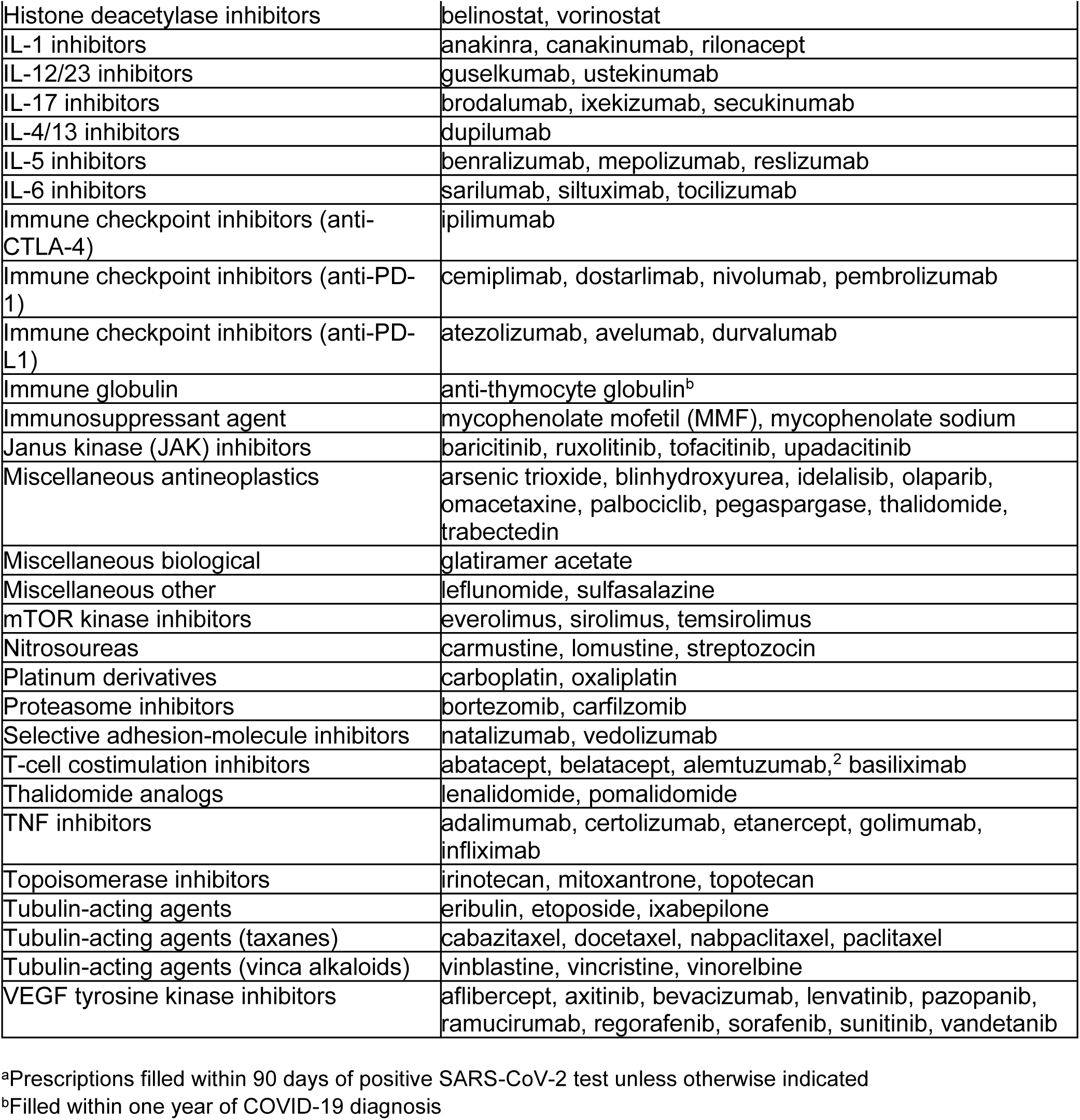
Immunosuppressive and cancer medications^a^

**Supplementary Table 3.**
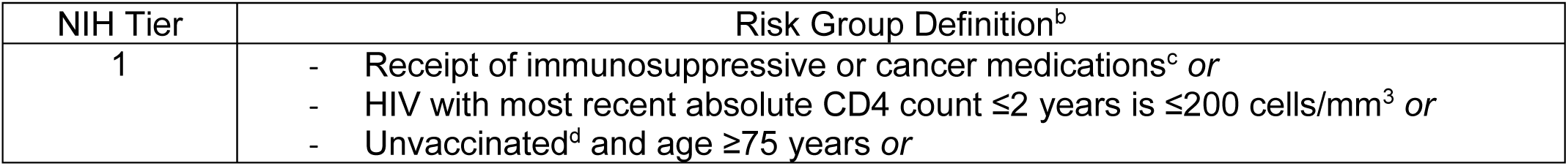
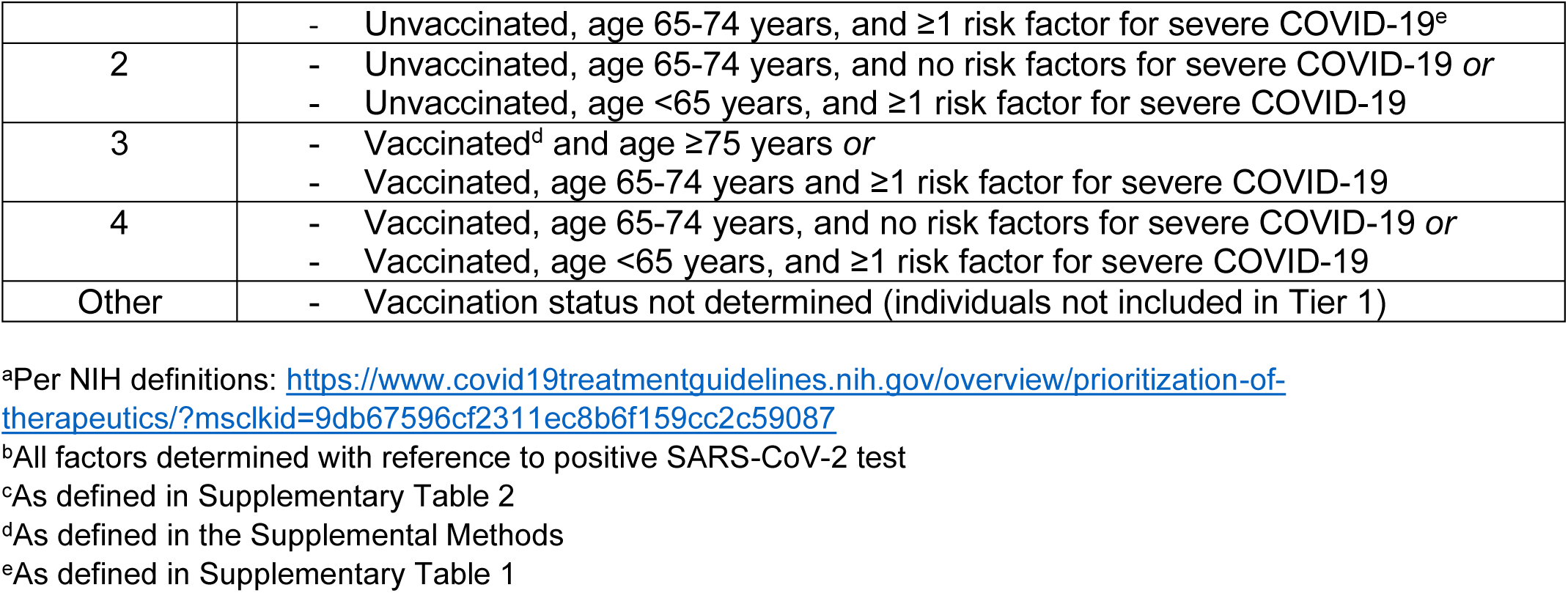
National Institutes of Health (NIH) tiers of prioritization for anti-SARS-CoV-2 therapies^a^

| NIH Tier | Risk Group Definition<sup>b</sup> |
| --- | --- |
| 1 | - Receipt of immunosuppressive or cancer medications<sup>c</sup> *or*<br>- HIV with most recent absolute CD4 count $\leq 2$ years is $\leq 200$ cells/mm<sup>3</sup> *or*<br>- Unvaccinated<sup>d</sup> and age $\geq 75$ years *or* |
| | - Unvaccinated, age 65-74 years, and $\geq 1$ risk factor for severe COVID-19<sup>e</sup> |
| 2 | - Unvaccinated, age 65-74 years, and no risk factors for severe COVID-19 *or*<br>- Unvaccinated, age <65 years, and $\geq 1$ risk factor for severe COVID-19 |
| 3 | - Vaccinated<sup>d</sup> and age $\geq 75$ years *or*<br>- Vaccinated, age 65-74 years and $\geq 1$ risk factor for severe COVID-19 |
| 4 | - Vaccinated, age 65-74 years, and no risk factors for severe COVID-19 *or*<br>- Vaccinated, age <65 years, and $\geq 1$ risk factor for severe COVID-19 |
| Other | - Vaccination status not determined (individuals not included in Tier 1) |

<sup>a</sup>Per NIH definitions: https://www.covid19treatmentguidelines.nih.gov/overview/prioritization-of-therapeutics/?msclkid=9db67596cf2311ec8b6f159cc2c59087
<sup>b</sup>All factors determined with reference to positive SARS-CoV-2 test
<sup>c</sup>As defined in Supplementary Table 2
<sup>d</sup>As defined in the Supplemental Methods
<sup>e</sup>As defined in Supplementary Table 1

**Supplementary Table 4.** Factors associated with receipt of different COVID-19 pharmacotherapies among Veterans, January-February 2022^a^

| Characteristic | Sotrovimab/<br>total<br>n/N (%) | Sotrovimab<br>Adjusted OR<br>(95% CI) | Nirmatrelvir/<br>total<br>n/N (%) | Nirmatrelvir<br>Adjusted OR<br>(95% CI) | Molnupiravir/<br>total<br>n/N (%) | Molnupiravir<br>Adjusted OR<br>(95% CI) | Remdesivir/<br>total n/N (%) | Remdesivir<br>Adjusted OR<br>(95% CI) |
| --- | --- | --- | --- | --- | --- | --- | --- | --- |
| Age, years |  |  |  |  |  |  |  |  |
| 18-49 | 80/28,890<br>(0.3) | 0.46 (0.35-0.6) | 265/28,890<br>(0.9) | 0.6 (0.51-0.69) | 104/28,890<br>(0.4) | 0.45 (0.36-0.57) | 58/28,890<br>(0.2) | 0.39 (0.29-0.53) |
| 50-64 (ref) | 176/30,749<br>(0.6) | - | 456/30,749<br>(1.5) | - | 239/30,749<br>(0.8) | - | 159/30,749<br>(0.5) | - |
| 65-74 | 382/24,815<br>(1.5) | 2.66 (2.22-3.19) | 530/24,815<br>(2.1) | 1.42 (1.25-1.61) | 300/24,815<br>(1.2) | 1.53 (1.28-1.81) | 189/24,815<br>(0.8) | 1.44 (1.17-1.79) |
| ≥75 | 273/16,133<br>(1.7) | 2.82 (2.32-3.43) | 326/16,133<br>(2.0) | 1.29 (1.12-1.5) | 214/16,133<br>(1.3) | 1.62 (1.34-1.95) | 139/16,133<br>(0.9) | 1.6 (1.26-2.02) |
| Female sex | 73/13,526<br>(0.5) | 1.03 (0.81-1.33) | 164/13,526<br>(1.2) | 0.99 (0.84-1.17) | 81/13,526<br>(0.6) | 1 (0.79-1.27) | 41/13,526<br>(0.3) | 0.77 (0.55-1.06) |
| Race |  |  |  |  |  |  |  |  |
| White (ref) | 753/74,093<br>(1.0) | - | 1,295/74,093<br>(1.7) | - | 703/74,093<br>(0.9) | - | 428/74,093<br>(0.6) | - |
| Black | 124/22,756<br>(0.5) | 0.61 (0.5-0.74) | 245/22,756<br>(1.1) | 0.63 (0.55-0.72) | 135/22,756<br>(0.6) | 0.65 (0.53-0.78) | 102/22,756<br>(0.4) | 0.83 (0.66-1.03) |
| Other | 34/3,738<br>(0.9) | 1.13 (0.8-1.59) | 37/3,738<br>(1.0) | 0.62 (0.45-0.87) | 19/3,738<br>(0.5) | 0.63 (0.4-0.99) | 15/3,738<br>(0.4) | 0.83 (0.49-1.39) |
| Hispanic<br>ethnicity | 61/8,609<br>(0.7) | 0.88 (0.67-1.14) | 127/8,609<br>(1.5) | 0.97 (0.81-1.17) | 47/8,609<br>(0.5) | 0.68 (0.51-0.92) | 37/8,609<br>(0.4) | 0.89 (0.64-1.25) |
| Rural residence | 204/16,137<br>(1.3) | 1.31 (1.12-1.55) | 298/16,137<br>(1.8) | 1.09 (0.96-1.25) | 190/16,137<br>(1.2) | 1.29 (1.1-1.53) | 99/16,137<br>(0.6) | 1.08 (0.86-1.35) |
| Region |  |  |  |  |  |  |  |  |
| West (ref) | 183/23,959<br>(0.8) | - | 322/23,959<br>(1.3) | - | 211/23,959<br>(0.9) | - | 106/23,959<br>(0.4) | - |
| Midwest | 241/18,831<br>(1.3) | 1.73 (1.41-2.11) | 395/18,831<br>(2.1) | 1.55 (1.33-1.81) | 196/18,831<br>(1.0) | 1.07 (0.88-1.31) | 40/18,831<br>(0.2) | 0.51 (0.35-0.74) |
| Northeast | 191/14,147<br>(1.4) | 1.84 (1.48-2.27) | 317/14,147<br>(2.2) | 1.71 (1.46-2.02) | 88/14,147<br>(0.6) | 0.66 (0.51-0.86) | 136/14,147<br>(1.0) | 2.3 (1.76-3.01) |
| South | 296/43,650<br>(0.7) | 0.99 (0.82-1.2) | 543/43,650<br>(1.2) | 0.95 (0.82-1.1) | 362/43,650<br>(0.8) | 0.95 (0.8-1.13) | 263/43,650<br>(0.6) | 1.47 (1.16-1.88) |
| COVID-19<br>vaccination |  |  |  |  |  |  |  |  |
| None (ref) | 245/30,671<br>(0.8) | - | 457/30,671<br>(1.5) | - | 196/30,671<br>(0.6) | - | 147/30,671<br>(0.5) | - |
| Partial | 61/3,917<br>(1.6) | 1.81 (1.35-2.43) | 77/3,917<br>(2.0) | 1.29 (1.01-1.67) | 35/3,917<br>(0.9) | 1.3 (0.89-1.9) | 36/3,917<br>(0.9) | 1.85 (1.27-2.68) |
| Full | 371/41,418<br>(0.9) | 0.98 (0.83-1.16) | 572/41,418<br>(1.4) | 0.87 (0.77-0.99) | 336/41,418<br>(0.8) | 1.19 (0.99-1.42) | 197/41,418<br>(0.5) | 0.88 (0.71-1.1) |
| Boosted | 234/24,557<br>(1.0) | 0.79 (0.65-0.95) | 471/24,557<br>(1.9) | 1.08 (0.94-1.24) | 290/24,557<br>(1.2) | 1.47 (1.21-1.78) | 165/24,557<br>(0.7) | 0.99 (0.78-1.25) |
| Smoking |  |  |  |  |  |  |  |  |
| Never (ref) | 327/39,061<br>(0.8) | - | 622/39,061<br>(1.6) | - | 306/39,061<br>(0.8) | - | 207/39,061<br>(0.5) | - |
| Former | 446/40,274<br>(1.1) | 1.09 (0.94-1.26) | 664/40,274<br>(1.6) | 0.92 (0.82-1.03) | 380/40,274<br>(0.9) | 1.04 (0.89-1.21) | 238/40,274<br>(0.6) | 0.98 (0.81-1.19) |
| Current | 102/17,286<br>(0.6) | 0.78 (0.62-0.97) | 236/17,286<br>(1.4) | 0.87 (0.75-1.01) | 149/17,286<br>(0.9) | 1.15 (0.94-1.4) | 75/17,286<br>(0.4) | 0.87 (0.67-1.14) |
| Alcohol<br>dependence | 137/22,658<br>(0.6) | 0.76 (0.63-0.91) | 273/22,658<br>(1.2) | 0.79 (0.69-0.9) | 145/22,658<br>(0.6) | 0.78 (0.65-0.94) | 91/22,658<br>(0.4) | 0.77 (0.61-0.98) |
| Substance<br>dependence | 23/5,400<br>(0.4) | 0.64 (0.42-0.99) | 56/5,400 (1) | 0.77 (0.59-1.01) | 40/5,400<br>(0.7) | 1.07 (0.77-1.48) | 35/5,400<br>(0.6) | 1.31 (0.9-1.91) |
| Number of<br>underlying<br>conditions |  |  |  |  |  |  |  |  |
| 1-2 (ref) | 108/37,502<br>(0.3) | - | 425/37,502<br>(1.1) | - | 167/37,502<br>(0.4) | - | 90/37,502<br>(0.2) | - |
| 3-4 | 255/31,289<br>(0.8) | 2.47 (1.95-3.13) | 534/31,289<br>(1.7) | 1.38 (1.21-1.58) | 277/31,289<br>(0.9) | 1.69 (1.38-2.06) | 156/31,289<br>(0.5) | 1.82 (1.38-2.4) |
| ≥5 | 543/29,852<br>(1.8) | 4.2 (3.34-5.29) | 599/29,852<br>(2.0) | 1.41 (1.23-1.63) | 410/29,852<br>(1.4) | 2.25 (1.85-2.75) | 293/29,852<br>(1.0) | 3.16 (2.42-4.13) |

<sup>a</sup>Results of multinomial and binomial models are similar. Multinomial models adjusted for age, sex, race, and ethnicity are presented and limited to Veterans with complete data for all included covariates

**Supplementary Table 5.** Factors associated with receipt of any COVID-19 pharmacotherapy among symptomatic Veterans, January-February 2022^a^

| Characteristic | Any treatment/total<br>n/N (%)<sup>b</sup> | Any treatment<br>Adjusted OR<br>(95% CI)<sup>c</sup> |
| --- | --- | --- |
| Age, years |  |  |
| 18-49 | 373/15,401 (2.4) | 0.48 (0.42-0.54) |
| 50-64 (ref) | 768/15,729 (4.9) | - |
| 65-74 | 1,009/12,221 (8.3) | 1.68 (1.53-1.86) |
| ≥75 | 692/7,581 (9.1) | 1.80 (1.62-2.01) |
| Female sex | 273/7,339 (3.7) | 0.63 (0.57-0.70) |
| Race |  |  |
| White (ref) | 2,308/37,204 (6.2) | - |
| Black | 453/11,860 (3.8) | 0.63 (0.57-0.70) |
| Other | 81/1,868 (4.3) | 0.83 (0.66-1.04) |
| Hispanic ethnicity | 184/4,438 (4.1) | 0.78 (0.67-0.91) |
| Rural residence<sup>a</sup> | 575/7,529 (7.6) | 1.31 (1.19-1.44) |
| Region |  |  |
| West (ref) | 606/12,216 (5.0) | - |
| Midwest | 632/9,829 (6.4) | 1.29 (1.15-1.45) |
| Northeast | 499/6,175 (8.1) | 1.64 (1.45-1.86) |
| South | 1,105/22,712 (4.9) | 1.04 (0.94-1.16) |
| COVID-19 vaccination<sup>b</sup> |  |  |
| None (ref) | 796/15,918 (5.0) | - |
| Partial | 168/2,278 (7.4) | 1.38 (1.16-1.65) |
| Full | 1,079/20,998 (5.1) | 0.93 (0.85-1.03) |
| Boosted | 799/11,726 (6.8) | 1.01 (0.91-1.12) |
| Smoking<sup>a</sup> |  |  |
| Never (ref) | 1,058/20,025 (5.3) | - |
| Former | 1,250/20,087 (6.2) | 1.01 (0.92-1.10) |
| Current | 432/8,914 (4.8) | 0.95 (0.84-1.07) |
| Alcohol dependence<sup>a</sup> | 462/1,1437 (4.0) | 0.77 (0.69-0.85) |
| Substance dependence<sup>c</sup> | 120/2,971 (4.0) | 0.86 (0.71-1.04) |
| Number of underlying<br>conditions<sup>a</sup> |  |  |
| 1-2 (ref) | 560/18,157 (3.1) | - |
| 3-4 | 870/15,696 (5.5) | 1.54 (1.38-1.72) |
| ≥5 | 1,391/16,195 (8.6) | 1.94 (1.74-2.17) |
| Cancer | 823/9,845 (8.4) | 1.32 (1.21-1.44) |
| Cardiovascular disease | 1,349/16,376 (8.2) | 1.39 (1.28-1.50) |
| Chronic kidney disease | 595/6,688 (8.9) | 1.35 (1.23-1.49) |
| Chronic lung disease | 1,171/16,035 (7.3) | 1.25 (1.16-1.36) |
| Diabetes | 1,217/15,697 (7.8) | 1.35 (1.25-1.46) |
| Immunosuppressive or<br>cancer medications<sup>d</sup> | 392/3,029 (12.9) | 2.47 (2.20-2.78) |
| Mental health conditions<sup>e</sup> | 1,115/22,342 (5.0) | 1.05 (0.97-1.14) |
| Obese (body mass index $\geq 30$ kg/m<sup>2</sup>) | 1,466/26,363 (5.6) | 1.16 (1.08-1.26) |

<sup>a</sup>Includes sotrovimab, nirmatrelvir, molnupiravir, and remdesivir. Symptomatic Veterans had any of 15 pre-specified COVID-19-related symptoms present on the day of positive SARS-CoV-2 test or within the preceding 30 days.
<sup>b</sup>A total of 3,079 Veterans who received any COVID-19 pharmacotherapy out of 56,206 Veterans testing positive for SARS-CoV-2 were included. Models were limited to Veterans with complete data for all included covariates.
<sup>c</sup>All models adjusted for age, sex, race, and ethnicity.

